# The Nicaraguan Pediatric Influenza Cohort Study, 2011-2019: influenza incidence, seasonality, and transmission

**DOI:** 10.1101/2022.02.01.22270201

**Authors:** Hannah E. Maier, Guillermina Kuan, Lionel Gresh, Gerardo Chowell, Kevin Bakker, Roger Lopez, Nery Sanchez, Brenda Lopez, Amy Schiller, Sergio Ojeda, Eva Harris, Angel Balmaseda, Aubree Gordon

## Abstract

**Background:** Children account for a large portion of global influenza burden and transmission, and a better understanding of influenza in children is needed to improve prevention and control strategies.

**Methods:** To examine the incidence and transmission of influenza we conducted a prospective community-based study of children aged 0-14 years in Managua, Nicaragua between 2011 and 2019. Participants were provided with medical care through study physicians and symptomatic influenza was confirmed by RT-PCR. Wavelet analyses were used to examine seasonality. Generalized growth models (GGMs) were used to estimate effective reproduction numbers.

**Results:** From 2011-2019, 3,016 children participated, with an average of ∼1,800 participants per year and median follow-up time of 5 years per child, and 48.3% of the cohort in 2019 had been enrolled their entire lives. The overall incidence rates per 100 person-years were 14.5 symptomatic influenza cases (95%CI: 13.9-15.1) and 1.0 influenza-associated ALRI case (95%CI: 0.8-1.1). Symptomatic influenza incidence peaked at age 9-11 months. Infants born during peak influenza circulation had lower incidence in the first year of their lives. The mean effective reproduction number was 1.2 (range 1.02-1.49), and we observed significant annual patterns for influenza and influenza A, and a 2.5-year period for influenza B.

**Conclusions:** This study provides important information for understanding influenza epidemiology and informing influenza vaccine policy. These results will aid in informing strategies to reduce the burden of influenza.

**Summary:** In this long-running influenza cohort, we found a substantial incidence of RT-PCR-confirmed influenza, and report by subtype/lineage. Infants born during influenza epidemics were protected from infection that first year. The mean effective reproduction number across years was 1.2.

---

Children make up a substantial portion of the considerable annual influenza burden [1, 2], especially in low-and middle-income countries [3]. Estimates of influenza burden vary greatly, in part because of poor data availability, and because most are not subtype and lineage-specific [4]. This lack of data is especially acute in low- and middle-income countries were currently ∼40% of the world’s population resides [5]. Children also contribute importantly to influenza transmission in communities [6], as shown by lower transmission during school closures [7] and with vaccination of children [8].

Early life influenza exposures are critical to the development of influenza immunity; it has been known for many years that prior influenza virus exposure impacts response to future strains [9-15]. In addition initial influenza exposures may provide protection against future pandemics of novel subtypes [14]. Understanding how early life exposures impact immunological response has major implications for public health, but requires that intensive long-term surveillance. The Nicaraguan Pediatric Influenza Cohort is the longest running influenza cohort that enrolls children from birth and is poised to provide critical data to aid in this understanding.

To optimize influenza prevention and control measures, particularly in resource-limited settings, a more accurate and thorough understanding of influenza burden and transmission is needed, especially in children. Here we present influenza incidence, seasonality and transmission estimates across a nine year period, in a long-running, urban community-based prospective cohort study of children in Nicaragua, a tropical lower-middle income country.

## METHODS

### Ethics Statement

Parent or guardian consent was obtained for all participants. In addition, verbal assent was obtained for participants aged six years and older. No monetary incentives were provided, but a small gift was provided for yearly for participation. The study was approved by the institutional review boards at the Nicaraguan Ministry of Health, the University of Michigan, and the University of California, Berkeley.

### Study Population

A detailed report on the design and methods of this cohort, including characteristics of the population, has been published previously [16]. Briefly, the Nicaraguan Pediatric Influenza Cohort Study (NPICS) is an ongoing prospective study (initiated in 2011) of children aged 0-14 years. The study is conducted in District II of Managua, Nicaragua, at the Health Center Sócrates Flores Vivas (HCSFV), and molecular testing for influenza is performed at the National Virology Laboratory (NVL) of the Centro Nacional de Diagnóstico y Referencia (CNDR). Initial enrollment in 2011 was done through random sampling of children aged 3-11 years enrolled in a prior cohort study of influenza [17, 18]; additional children aged 0-2 years were recruited through house-to-house visits. Newborn infants ≤4 weeks are enrolled monthly into NPICS. Children may remain in the study until their 15^th^ birthday. All participants live within the catchment area of the HCSFV.

### Follow-up and case identification

Once enrolled, children are followed with annual surveys, and families are asked to bring their children to the HCSFV for medical care at the first sign of illness (study diagram in appendix p3). The criteria for influenza testing by real-time reverse-transcription polymerase chain reaction (RT-PCR) was measured or reported fever (37.8C) or feverishness with cough, sore throat, or runny nose for children aged 2 years and older, and only fever or feverishness for children under 2. Primary medical care and laboratory testing are provided to all participants, free of charge. Multiple attempts are made to reach the participants, including house visits, before considering them lost to follow-up. The primary outcome in this analysis is symptomatic RT-PCR-confirmed influenza infection. A secondary outcome is influenza-associated acute lower respiratory infection (ALRI), defined as pneumonia, bronchitis, bronchiolitis, bronchopneumonia, or bronchial hyper-reactivity occurring up to fourteen days prior to and 28 days after a positive influenza RT-PCR test. Some children were also enrolled in influenza household transmission studies starting in 2012, which tested for influenza regardless of symptoms; influenza-positive RT-PCR tests from these children who did not meet the testing criteria were not included in this analysis. Six un-subtyped influenza A cases in 2017 and 2019 were re-coded as H3N2, as that was the nearly exclusive circulating subtype those years. Lineage was not available for 50 influenza B infections.

### Statistical Analysis

#### Incidence

Person-time was calculated as the amount of time between enrollment and December 31, 2019, or withdrawal. For those lost to follow-up, exit dates were calculated as the midpoint between the date of last contact and the date recorded by study personnel as lost to follow-up. Incidence and confidence intervals were calculated using intercept-only generalized linear models with Poisson distributions. Analyses were performed for all influenza, by type (A and B), by influenza A subtype (H1N1pdm and H3N2), and by influenza B lineage (Victoria and Yamagata). Participant age used in incidence models was calculated on a weekly basis. R version 4.1.0 was used to perform analyses and create figures [19].

#### Effective Reproduction Numbers

We used generalized growth models (GGMs) [20] to estimate the effective reproduction numbers of yearly influenza epidemics. GGMs allow sub-exponential epidemic growth early on, by linking the serial interval (assumed here to be 2.85 (standard deviation 0.93) days [21, 22]) with case incidence time series and estimating the transmission potential over time. The GGMs additionally estimate two more parameters, the epidemic growth rate and a scaling factor for the epidemic growth rate, which can range from zero (no growth) to one (exponential epidemic growth). We used Wilcoxon tests to compare parameter values between influenza (sub)types.

#### Seasonality

To analyze seasonal patterns, in addition to plotting weekly influenza incidence, wavelet analyses using pink noise were performed to identify dominant and significant seasonal cycles [23]. Data were analyzed using a log+1 transformation to better interpret the seasonal patterns.

## RESULTS

### Study Participation

Between 2011 and 2019, 3,016 children were enrolled, contributing a total of 16,352 person-years (table 1), with an average of ∼1,800 participants enrolled per year and a median follow-up time of 5 years per child (IQR: 3-8 years, figure 1A,D,E). Overall, 35.7% of the cohort was enrolled for their whole lives (enrolled in their first 3 months of life), and by 2019, 48.3% of active participants in 2019 had been enrolled their whole lives, because most enrollees after 2011 were newborns (figure 1B). A total of 607 children (20.1%) completed the study (most aged out but four moved out of the study catchment area), 544 children (18.0%) were ultimately lost to follow-up, 132 (4.4%) were ultimately withdrawn, and 7 (0.2%) died (figure 1C); 1,726 children (57.2%) were still enrolled at the end of 2019. After being lost or withdrawn, eight children re-enrolled; one completed the study; two were lost again, and five were still enrolled at the end of 2019. The mean yearly loss to follow-up was 3.4% (range 1.9-5.7%, figure 1C). Six of the seven deaths were in children under one year of age, and the other death was a ten-year-old in 2016. This population has very low rates of influenza vaccination, with 13.3% ever vaccinated and 2.7% vaccinated yearly (rates by age and year are presented in appendix p 4). Children presented at the health center 71,123 times, of which 17,531 (24.6%) visits met the study testing definition for influenza. Of these visits, 16,439 (93.8%) presented for care within four days of onset of symptoms.

**Table 1.** Participant Characteristics

|  | Person-years* |  | Participants |
| --- | --- | --- | --- |
|  | N | % |  |
| Total | 16352 | 100.0 | 3016 |
| Age |  |  |  |
| <1y | 1035 | 6.3 |  |
| 1-4y | 4421 | 27.0 |  |
| 5-10y | 6409 | 39.2 |  |
| 11-12y | 2175 | 13.3 |  |
| 13-14y | 2312 | 14.1 |  |
| Sex |  |  |  |
| F | 8231 | 50.3 | 1513 |
| M | 8121 | 49.7 | 1503 |

\*counting every year partially enrolled

**Figure 1.**
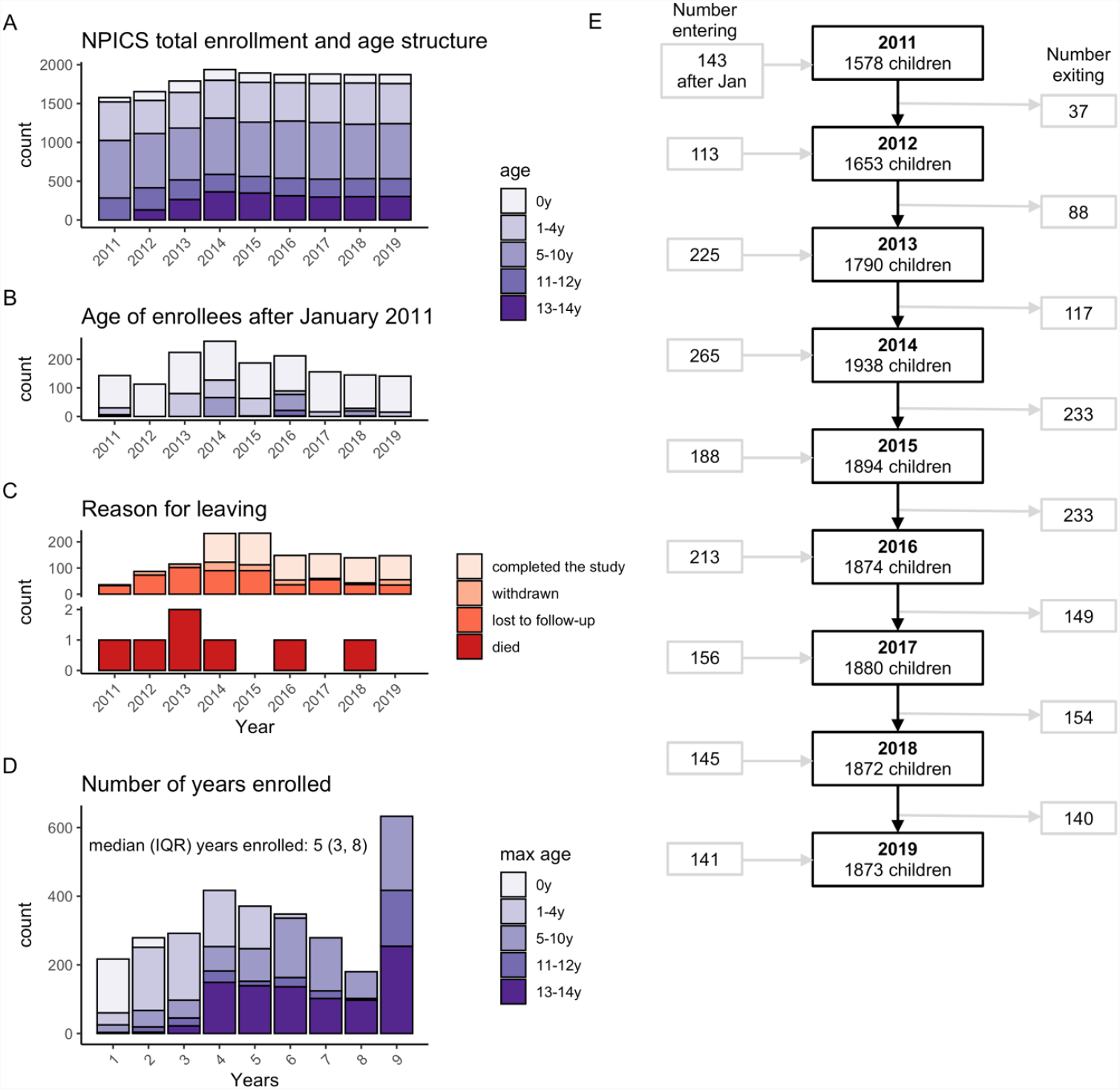
Characteristics of the Nicaraguan Influenza Pediatric Cohort (NPICS), by year, 2011-2019. A) Total enrollment over time by age, B) age of enrollees after the initial enrollment by year, C) reasons for leaving the cohort by year, D) number of years enrolled in the cohort by age, and E) flowchart of the number of participants by year. Note that some children exited and then re-entered in the same year.

### Influenza Cases

Between 2011 and 2019, 2,176 suspected infections were RT-PCR-confirmed as influenza (table 2). Close to half of participants (1,411 [46.8%]) experienced at least one symptomatic influenza infection while enrolled; 36% ever had an influenza A infection (18% ever had H1N1pdm, 26% ever had H3N2) and 23% ever had an influenza B infection (12% ever had Victoria, and 12% ever had Yamagata), with up to 5 total influenza illnesses per child. There were two co-infections, both with H1N1pdm and H3N2 in 2011. The overall incidence of symptomatic influenza was 14.5 cases (95%CI: 13.9-15.1) and 1.0 influenza-associated ALRI case (0.8-1.2) per 100 person-years (table 2). Among children with two or more influenza illnesses, the mean time between illnesses was 2.0 years with a range of one week (different influenza types) to almost nine years. The mean time between influenza illnesses increased with age (p<0.0001), from 1.3 years for children aged 0-4 years to 2.4 years for children aged 5-14 years (appendix p 5).

**Table 2.** Incidence of symptomatic influenza and influenza-associated ALRI in the Nicaraguan Pediatric Influenza Cohort Study (NPICS), 2011-2019

|  | PY | All Influenza |  | Influenza A |  | Influenza B |  | H1N1pdm |  | H3N2 |  | Victoria |  | Yamagata |  |
| --- | --- | --- | --- | --- | --- | --- | --- | --- | --- | --- | --- | --- | --- | --- | --- |
|  |  | N | Inc (CI) | N | Inc (CI) | N | Inc (CI) | N | Inc (CI) | N | Inc (CI) | N | Inc (CI) | N | Inc (CI) |
| Symptomatic Influenza |  |  |  |  |  |  |  |  |  |  |  |  |  |  |  |
| Everyone | 14982.0 | 2176 | 14.5 (13.9, 15.1) | 1403 | 9.4 (8.9, 9.9) | 773 | 5.2 (4.8, 5.5) | 541 | 3.6 (3.3, 3.9) | 864 | 5.8 (5.4, 6.2) | 368 | 2.5 (2.2, 2.7) | 355 | 2.4 (2.1, 2.6) |
| Age |  |  |  |  |  |  |  |  |  |  |  |  |  |  |  |
| 0-4y | 5176.6 | 1046 | 20.2 (19.0, 21.5) | 752 | 14.5 (13.5, 15.6) | 294 | 5.7 (5.1, 6.4) | 286 | 5.5 (4.9, 6.2) | 466 | 9.0 (8.2, 9.9) | 145 | 2.8 (2.4, 3.3) | 130 | 2.5 (2.1, 3.0) |
| 5-10y | 6246.1 | 825 | 13.2 (12.3, 14.1) | 468 | 7.5 (6.8, 8.2) | 357 | 5.7 (5.2, 6.3) | 191 | 3.1 (2.7, 3.5) | 279 | 4.5 (4.0, 5.0) | 172 | 2.8 (2.4, 3.2) | 166 | 2.7 (2.3, 3.1) |
| 11-14y | 3559.3 | 305 | 8.6 (7.7, 9.6) | 183 | 5.1 (4.4, 5.9) | 122 | 3.4 (2.9, 4.1) | 64 | 1.8 (1.4, 2.3) | 119 | 3.3 (2.8, 4.0) | 51 | 1.4 (1.1, 1.9) | 59 | 1.7 (1.3, 2.1) |
| Sex |  |  |  |  |  |  |  |  |  |  |  |  |  |  |  |
| F | 7553.9 | 1085 | 14.4 (13.5, 15.2) | 706 | 9.3 (8.7, 10.1) | 379 | 5.0 (4.5, 5.5) | 266 | 3.5 (3.1, 4.0) | 441 | 5.8 (5.3, 6.4) | 181 | 2.4 (2.1, 2.8) | 178 | 2.4 (2.0, 2.7) |
| M | 7428.1 | 1091 | 14.7 (13.8, 15.6) | 697 | 9.4 (8.7, 10.1) | 394 | 5.3 (4.8, 5.9) | 275 | 3.7 (3.3, 4.2) | 423 | 5.7 (5.2, 6.3) | 187 | 2.5 (2.2, 2.9) | 177 | 2.4 (2.1, 2.8) |
| Influenza-associated ALRI |  |  |  |  |  |  |  |  |  |  |  |  |  |  |  |
| Everyone | 14982.0 | 147 | 1.0 (0.8, 1.2) | 110 | 0.7 (0.6, 0.9) | 37 | 0.2 (0.2, 0.3) | 44 | 0.3 (0.2, 0.4) | 66 | 0.4 (0.3, 0.6) | 20 | 0.1 (0.1, 0.2) | 11 | 0.1 (0.0, 0.1) |
| Age |  |  |  |  |  |  |  |  |  |  |  |  |  |  |  |
| 0-4y | 5176.6 | 111 | 2.1 (1.8, 2.6) | 88 | 1.7 (1.4, 2.1) | 23 | 0.4 (0.3, 0.7) | 37 | 0.7 (0.5, 1.0) | 51 | 1.0 (0.7, 1.3) | 16 | 0.3 (0.2, 0.5) | 4 | 0.1 (0.0, 0.2) |
| 5-10y | 6246.1 | 26 | 0.4 (0.3, 0.6) | 17 | 0.3 (0.2, 0.4) | 9 | 0.1 (0.1, 0.3) | 7 | 0.1 (0.1, 0.2) | 10 | 0.2 (0.1, 0.3) | 2 | 0.0 (0.0, 0.1) | 5 | 0.1 (0.0, 0.2) |
| 11-14y | 3559.3 | 10 | 0.3 (0.2, 0.5) | 5 | 0.1 (0.1, 0.3) | 5 | 0.1 (0.1, 0.3) | 0 | - | 5 | 0.1 (0.1, 0.3) | 2 | 0.1 (0.0, 0.2) | 2 | 0.1 (0.0, 0.2) |
| Sex |  |  |  |  |  |  |  |  |  |  |  |  |  |  |  |
| F | 7553.9 | 60 | 0.8 (0.6, 1.0) | 44 | 0.6 (0.4, 0.8) | 16 | 0.2 (0.1, 0.3) | 14 | 0.2 (0.1, 0.3) | 30 | 0.4 (0.3, 0.6) | 7 | 0.1 (0.0, 0.2) | 6 | 0.1 (0.0, 0.2) |
| M | 7428.1 | 87 | 1.2 (0.9, 1.4) | 66 | 0.9 (0.7, 1.1) | 21 | 0.3 (0.2, 0.4) | 30 | 0.4 (0.3, 0.6) | 36 | 0.5 (0.3, 0.7) | 13 | 0.2 (0.1, 0.3) | 5 | 0.1 (0.0, 0.2) |

PY= person-years, CI = 95% Confidence Interval. Incidence is expressed as cases per 100 person-years.

### Incidence Comparisons by Age and Sex

Children aged 9-11 months had the highest influenza burden, with 26.1 cases of symptomatic influenza (95%CI: 20.6-32.9) and 4.8 influenza-associated ALRI cases (95%CI: 2.8-8.3) per 100 person-years, increasing from 7.2 symptomatic influenza cases (95%CI: 4.3-11.9) and 1.0 influenza-associated ALRI case (95%CI: 0.2-3.8) in children aged <3 months and then decreasing by age 14 years to 7.8 symptomatic influenza cases (95%CI: 6.0-10.2) and 0.3 influenza-associated ALRI cases (95%CI: 0.1-1.2, figure 2). By age groups 0-4, 5-10, and 11-14 years, incidence of symptomatic influenza was 20.2 (95%CI: 19.0-21.5), 13.2 (95%CI: 12.3-14.1), and 8.6 (95%CI: 7.7-9.6), and incidence of influenza-associated ALRI was 2.1 (95%CI: 1.8-2.6), 0.4 (95%CI: 0.3-0.6), and 0.3 (95%CI: 0.2-0.5) per 100 person-years, respectively (table 2). The same age patterns were true of incidence for influenza A and both subtypes, but were somewhat different for influenza B, for which symptomatic influenza incidence was similar for ages 0-4 and 5-10 and lower by age 11-14, and influenza-associated ALRI incidence was higher for children aged 0-4 and similar for older ages (table 2). There were no significant differences by sex, but males had slightly more influenza-associated ALRI (M: 1.2, 95%CI: 0.9-1.4; F: 0.8, 95%CI: 0.6-1.0, (table 2).

**Figure 2.**
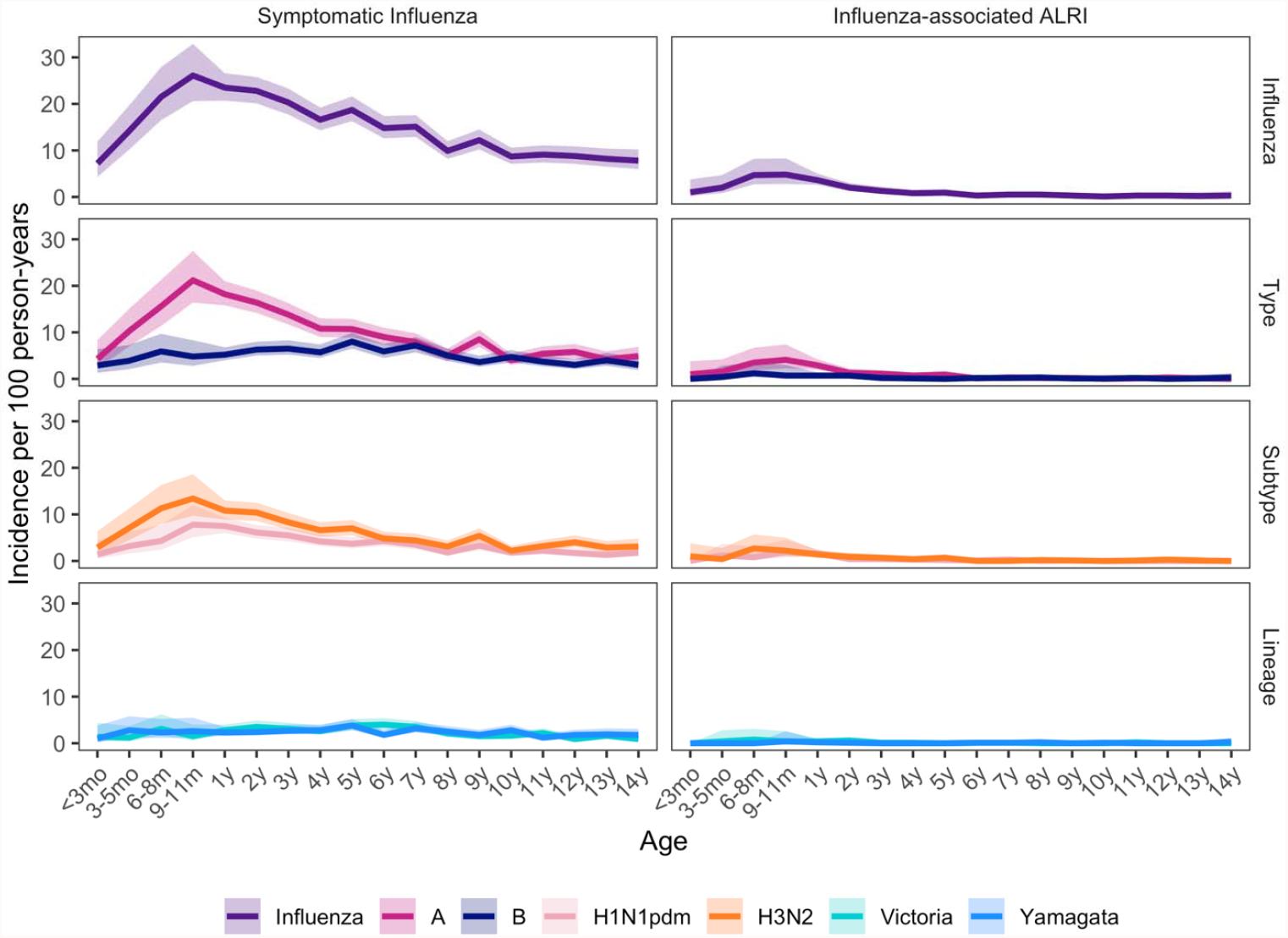
Incidence of symptomatic influenza and influenza-associated ALRI by age, for all influenza and by (sub)type and lineage. Age groups are every three months until age one year, then yearly thereafter. Shaded regions show 95% confidence intervals.

### Infant Immunity

We investigated immunity in children’s first and second influenza seasons by selecting infants who were enrolled in their first six months of life and comparing incidence of symptomatic influenza by month born and month of infection, across all study years (figure 3). In their first year enrolled, incidence was highest for children born in May (31 cases per 100 person-years, 95%CI: 18.7-51.5), while peak month of infection was 5 months later than their births, in October; incidence was lowest for children born in July (9.2 cases per 100 person-years, 95%CI: 3.0-28.6, figure 3A). Children born from December-June had 23.8 cases per 100 person-years (95%CI: 19.6-28.8), compared to only 13.9 cases per 100py in children born during the primary months of influenza circulation, July-November (95%CI: 9.2-20.9, figure 3B). This pattern was similar for influenza A and B in the first year enrolled, although for A this was primarily driven by H3N2 and there was no difference for H1N1pdm (figure 3B, appendix p 6).

**Figure 3.**
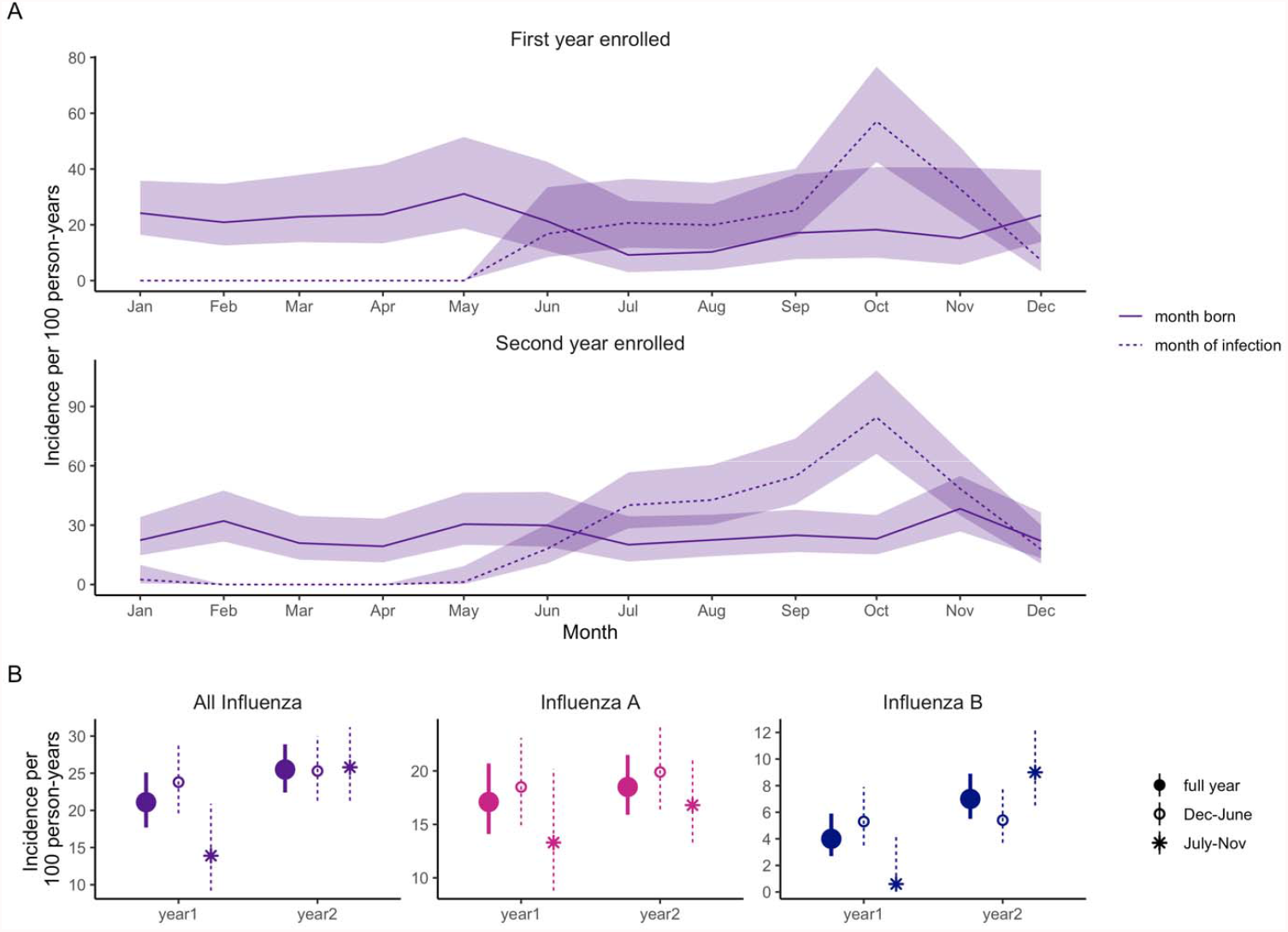
Symptomatic influenza incidence by month born. This figure only includes children enrolled in their first six months of life. Panel A shows symptomatic influenza incidence by month born (solid lines) and month of infection (dashed lines) in the first (top) and second (bottom) years enrolled. Panel B shows the overall symptomatic influenza incidence for the first two years enrolled (solid circles), and incidence categorized by month born (December-June births in open circles and July-November births in asterisks).

In their second year enrolled, this effect disappeared (figure 3A&B). Interestingly, for influenza B in the second year enrolled, there was a catch up in incidence for the children born during primary influenza circulation months July-November (figure 3B), with 9.0 vs 5.4 cases per 100py compared to children born December-June (95%CI: 6.5-12.4 vs 3.7-7.8).

### Effective Reproduction Numbers

The overall mean estimated effective reproduction number (R effective) was 1.20 (figure 4A), with a range in yearly estimates from 1.02 in 2012 to 1.49 in 2015 (figure 4B). R effective appeared marginally higher for influenza A vs B, however with a small number of seasons to compare (n = 9 and n = 6), we did not observe a statistically significant difference (A vs B: p = 0.224, H3N2 vs B: p = 0.101). The ascending period—the time in weeks from epidemic start to peak—ranged from 4 weeks in 2011 to 11 weeks in 2012 and was longest for influenza B (A vs B: p = 0.091, H3N2 vs B: p = 0.043, appendix p 7). The growth rate scaling parameter for influenza ranged from 0.26 in 2012 to 0.95 in 2016, indicating a range in growth behavior from sub-exponential to nearly exponential (appendix p 7).

**Figure 4.**
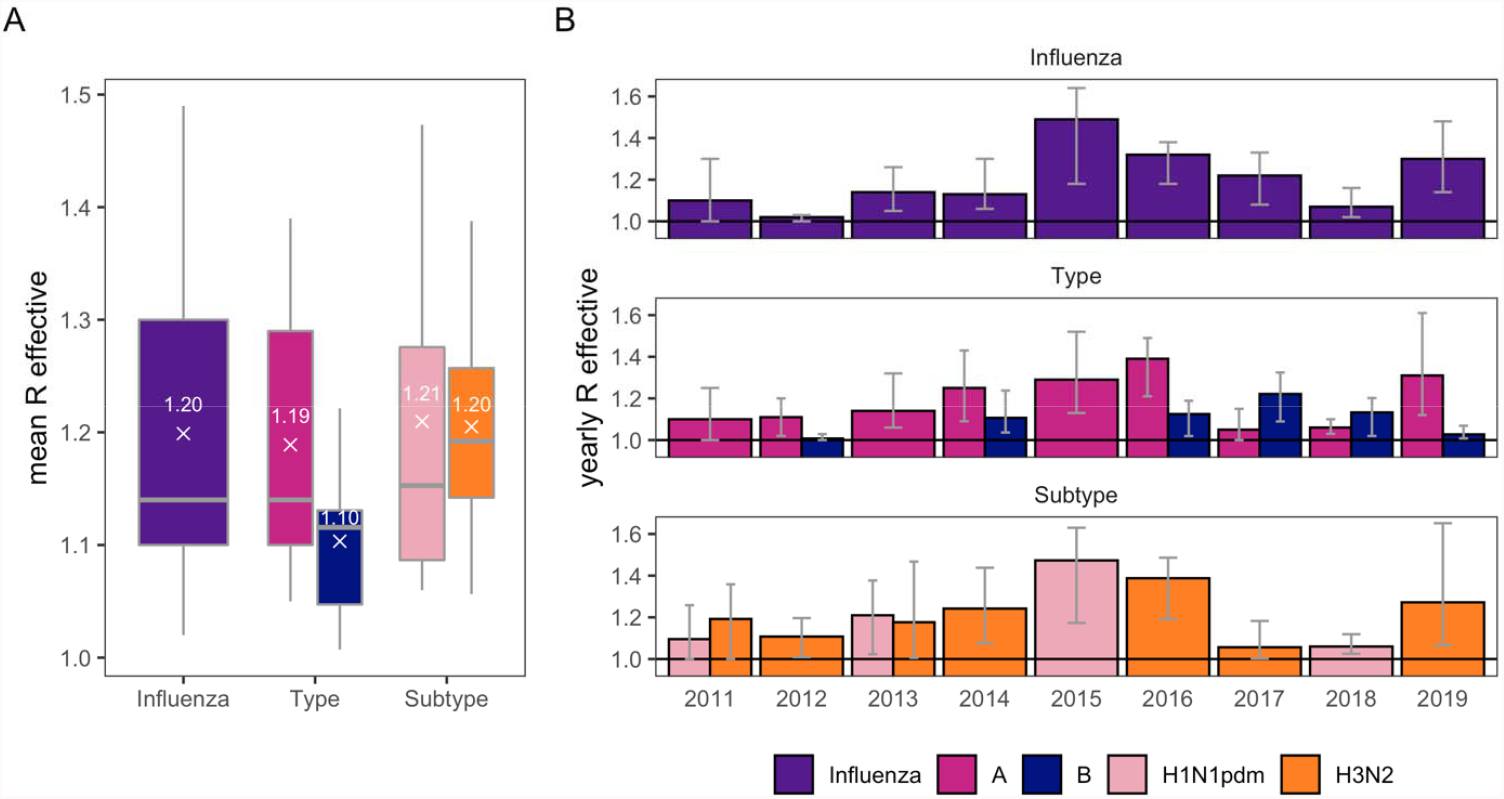
Effective reproduction numbers, for all influenza, by type (A and B), and by A subtype (H1N1pdm and H3N2). A) Mean effective reproduction numbers summarized across years. B) Effective reproduction numbers estimates and confidence intervals by year.

### Seasonality

Circulation of influenza subtypes and lineages alternated from 2011-2019 (figure 5A and appendix p 11), and 2007-2010 (data from a prior study[17], inset of figure 5A), with typically one epidemic per year unless there were large epidemics of multiple influenza types, as in 2010, 2014, and 2017, which occurred close together in time (June/August peaks in 2010, August/September peaks in 2014 and June/August peaks in 2017). Years with the highest number of symptomatic influenza cases did not correspond to the years with the highest influenza-associated ALRI cases: for example, 2013 with H3N2 predominating had an average incidence of symptomatic influenza but the highest incidence of influenza-associated ALRI (appendix p 8). Interestingly, 2014, which had an even higher incidence of symptomatic H3N2 cases, had lower H3N2-associated ALRI incidence (appendix p 8).

**Figure 5.**
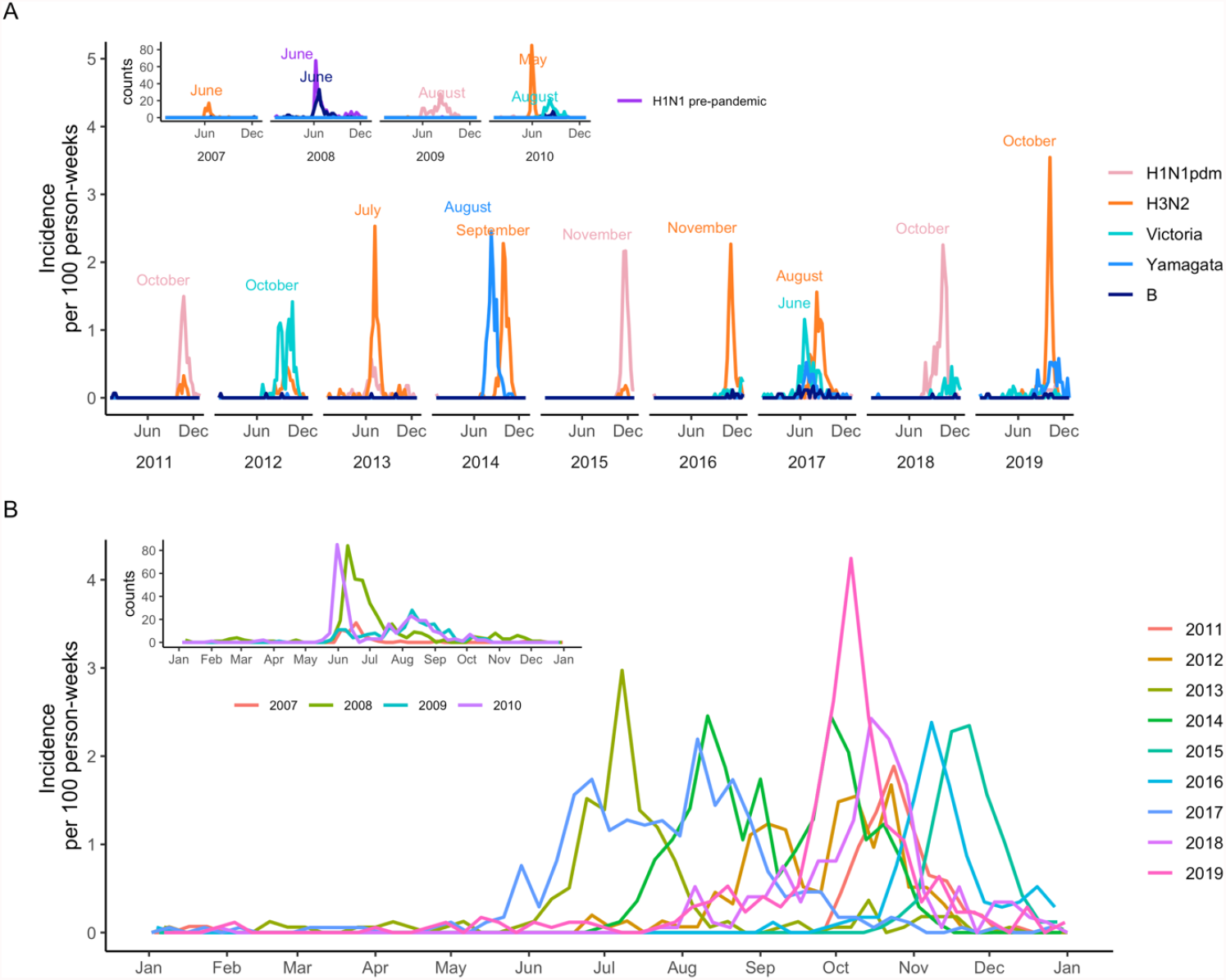
Symptomatic influenza incidence over time. A) Circulation of influenza subtypes and lineages over the study years 2011-2019. Lineage was not available for 50 influenza B infections. Peak months are listed. B) Yearly influenza epidemics by month (all subtypes and lineages combined). In both panels, insets show influenza counts from this study setting for years 2007-2010; data come from an earlier pilot study (described in Gordon et al. 2009).

Seasonal influenza epidemic peaks alternated between June-August and September-December, but nearly all (97.3%) circulation occurred from June-December (Figure 5A&B, appendix p9). From 2011-2019, influenza circulated later and with more September-December peaks, compared to the earlier 2007-2010 cohort (Figure 5A&B, appendix p9). Wavelet analysis confirmed a significant annual pattern (appendix p 10); similar patterns are seen for influenza A (appendix p 10). For influenza B, wavelet analysis showed a 2.5-year dominant period; this fits with the semi-alternating years of influenza B circulation (figure 5A) and two large relatively early seasons with peaks in August 2014 and June 2017 (appendix p 10).

## DISCUSSION

In this long-running prospective cohort study of influenza in children in Nicaragua, we found a significant burden of laboratory-confirmed symptomatic influenza. Children aged 9-11 months had the highest incidence, and incidence declined with age. Infants born during the primary months of influenza circulation had lower incidence compared to those born when influenza was not circulating. Across nine years, the overall mean effective reproduction number was 1.2, and one large annual epidemic occurred—sometimes one influenza type followed shortly by the other— with seasonal peaks alternating between June-August and September-December and nearly all circulation between June-December.

Compared to other community-based studies of multiple seasons, our reported childhood influenza burden (14.5 symptomatic influenza cases and 1.0 influenza-associated ALRI case per 100 person-years) was higher than in wealthier, more vaccinated settings of Michigan (10.7 and 9.1 influenza cases per 100 person-seasons in children 0-4y and 5-17, respectively [24]) and Finland (3.6 cases per 100 person years in children 0-2y [25]), and more comparable to (semi) tropical Peru (21.4, 25.6, 20.4, and 11.2 cases per 100 person years in children <2y, 2-4y, 5-11y, and 12-17y, respectively [26]) and Hong Kong (4-21 cases per 100 person-season over 4 seasons, in children 2-19y [27]). To our knowledge, ours is the largest, longest-running community-based prospective pediatric cohort study. Because influenza epidemics vary in (sub)type, lineage, size, and severity from year-to-year, studying only a few seasons will not capture the variability in incidence and burden, especially since immunity develops over time, strongly influenced by first exposures. However, estimates averaged over multiple seasons will provide an accurate picture. While data exists on the incidence of influenza in wealthy and temperate regions, detailed reporting of influenza in the tropics, Latin America, and LMICs is much poorer, particularly in children [28, 29].

Others have shown patterns of influenza immunity by birth month: children in the US aged 2-5 years were more likely to be vaccinated if their birthdays occurred between September and December, with those children also less likely to be diagnosed with influenza. This likely occurred because young children typically have their annual checkups near their birthday, meaning they’d be vaccinated [30]. Pregnant women are a priority group for influenza vaccination because of the protection afforded to newborns [31], and our finding that incidence was lower for infants born during influenza epidemics is likely related to passively acquired maternal immunity, which declines over the first 6-12 months of life [32]. Unfortunately, we do not have information on maternal vaccination status, but our results suggest that whether maternal antibodies were from infection or vaccination, they provided some protection to newborns. Protection among infants born during influenza epidemics and highest incidence among children aged 9-11 months highlight the importance in protecting infants of vaccinating pregnant women, postpartum women who breastfeed [33], and infants as soon as possible after the recommended age of six months.

Our estimate of the effective reproduction number for Nicaragua from 2011-2019 (1.2, range across years of 1.0-1.5) falls between estimates for seasonal influenza from 1996-2006 in Brazil, another (semi) tropical Latin American country, of 1.03 (95%CI: 1.02-1.04) [34], and a median estimate for seasonal influenza from 24 studies and many years going as far back as 1889, mostly from Israel, Europe, and North America, of 1.27 (IQR: 1.19-1.37) [35]. It was also below the median 2009 H1N1 pandemic estimate, summarized from 78 estimates from many locations, of 1.46 (IQR: 1.30-1.70) [35], but of note, our highest sub-type-specific R effective was for H1N1pdm in 2015 (1.5, 95% CI: 1.2-1.6).

Influenza has been thought to lack seasonality in the tropics [28, 29], however, we saw practically no influenza cases from January through May. This misconception may be due to the previous insufficient surveillance and reporting scale. For example, there were H1N1pdm peaks at the end of 2015, middle of 2016, and end of 2016 in Central America [29], but in Managua, we only saw H1N1pdm circulation at the end of 2015. It’s possible that local transmission is seasonally distinct in the tropics, but there are enough important small-scale geographic differences that seasonality is blurred when aggregated. Influenza seasonality in Nicaragua has changed from two annual peaks, primary in June/July and secondary in November/December, prior to the 2009 H1N1 pandemic [17, 36], to one annual peak after 2009, alternating between earlier and later timing. The optimal timing for influenza vaccination in Managua is likely in the several months after the Southern Hemisphere vaccine becomes available in April. Seasonal differences between Managua and Central America suggest that the timing of epidemics vary, and that optimal timing of vaccination may vary by region.

Our ongoing long-running community-based prospective pediatric cohort study has added important details to our understanding of influenza burden in children and fills a gap in information on influenza in the tropics and in Latin America in particular [28, 29]. These results provide much-needed information on influenza in a tropical LMIC setting and will be invaluable in informing strategies to reduce the burden of influenza.

## Data Availability

Individual-level data may be shared with outside investigators following University of Michigan IRB approval. Please contact Aubree Gordon to arrange for data access.

## FUNDING

This work was supported by the National Institute for Allergy and Infectious Diseases at the National Institute of Health [award no. R01 AI149747-01 and contract no. HHSN272201400006C to A.G., U01 AI44616 to A.G., U01 AI088654 to A.G. and E.H.].

## ACKNOWLEDGEMENTS

We are extremely grateful to the children who participated in this study and to their families, and to the incredibly dedicated teams at the Centro de Salud Sócrates Flores Vivas and the Nicaraguan National Virology Laboratory at the Nicaraguan Ministry of Health and at the Sustainable Sciences Institute.

## COMPETING INTERESTS

Aubree Gordon serves on an advisory board for Janssen. All other authors report no competing interests.

## Notes

### Author Declarations

The study was approved by the institutional review boards at the Nicaraguan Ministry of Health, the University of Michigan, and the University of California, Berkeley.

