## appendix for "The Nicaraguan Pediatric Influenza Cohort Study, 2011-2019: influenza incidence, seasonality, and transmission"

^1^Department of Epidemiology, School of Public Health, University of Michigan, Ann Arbor, Michigan, USA; ^2^Sustainable Sciences Institute, Managua, Nicaragua; ^3^Centro de Salud Sócrates Flores Vivas, Ministry of Health, Managua, Nicaragua; ^4^ Laboratorio Nacional de Virología, Centro Nacional de Diagnóstico y Referencia, Ministry of Health, Managua, Nicaragua; ^5^Division of Infectious Diseases and Vaccinology, School of Public Health, University of California, Berkeley; ^6^Population Health Sciences, School of Public Health, Georgia State University

* co-first authors^; $^ co-last authors

Table of Contents

Figure S1. NPICS study diagram…………………………………………………………………………………………….3

Figure S2. Influenza vaccination in the Nicaraguan Pediatric Influenza Cohort, by age and year…………..........................4

Figure S3. Time since last influenza episode, by age……………………………………………………………………......5

Figure S4. Symptomatic influenza incidence in the first and second years enrolled in infants enrolled in their first 6 months of life, by (sub)type and lineage……………………………………………………………………………………..6

Figure S5. Other estimated transmission parameters………………………………………………………………………...7

Figure S6. Incidence of symptomatic influenza and influenza-associated ALRI in the Nicaraguan Pediatric Influenza Cohort Study (NPICS), 2011-2019, by year…………………………….…………………………………………………...8

Figure S7. Influenza epidemic timing………………………………………………………………………………………..9

Figure S8. Influenza seasonality by Wavelet analysis……………………………………………………….......................10

Figure S9. Yearly influenza epidemics, by month………………………………………………………………………….11

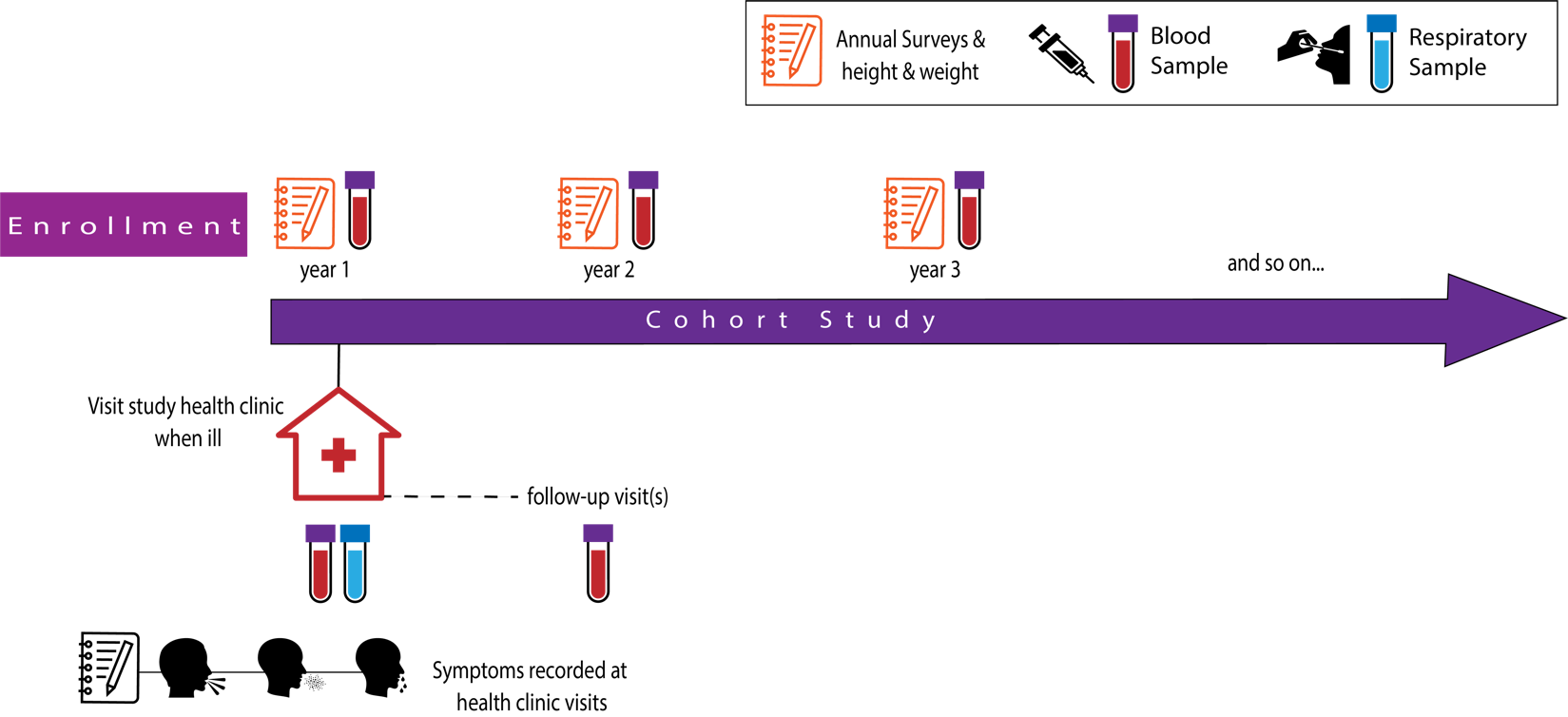

Figure S1. NPICS study diagram.

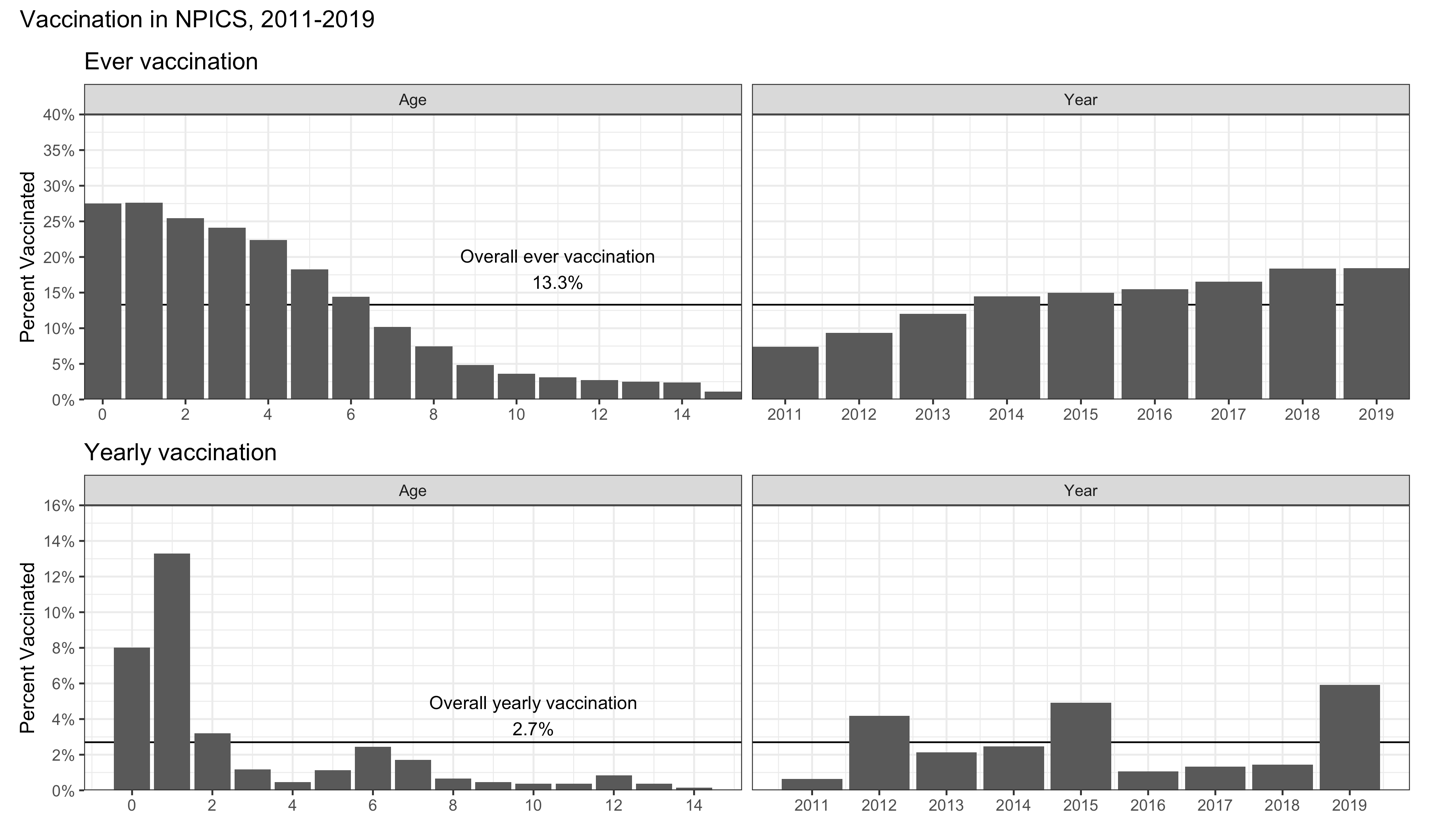

Figure S2. Influenza vaccination in the Nicaraguan Pediatric Influenza Cohort, by age (on March 1) and year. Vaccination was defined as at least one vaccine dose in the year.

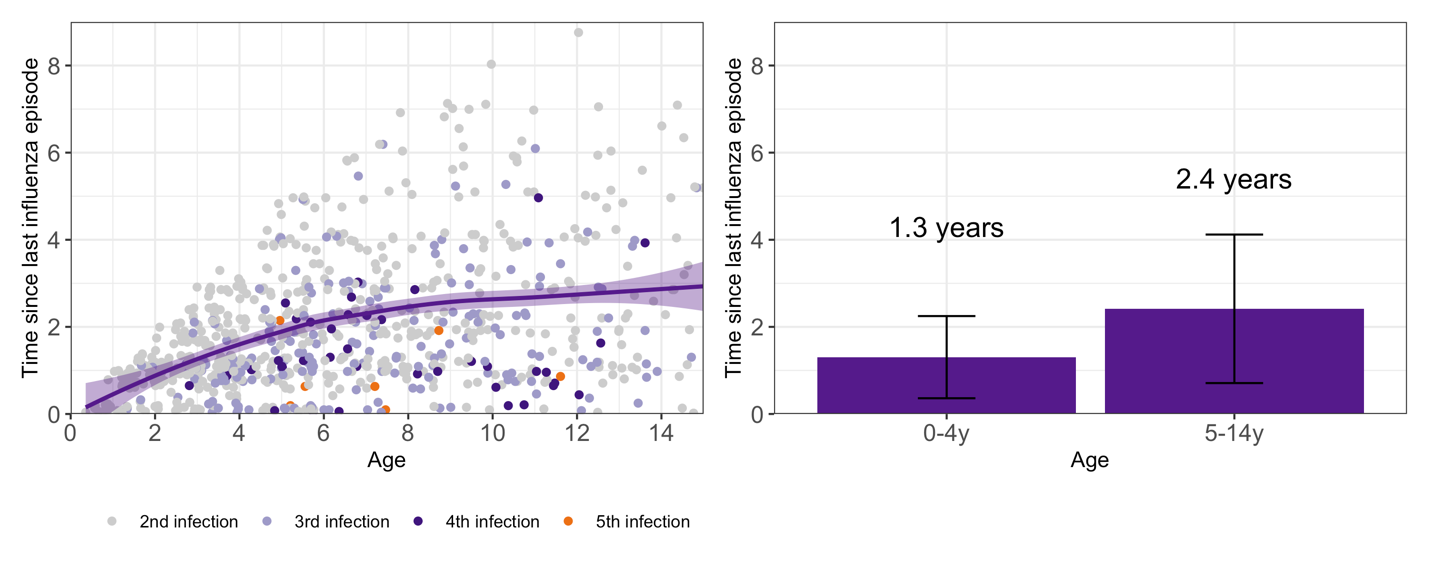

Figure S3. Time since last influenza episode, by age. Among children with at least 2 symptomatic influenza infections.

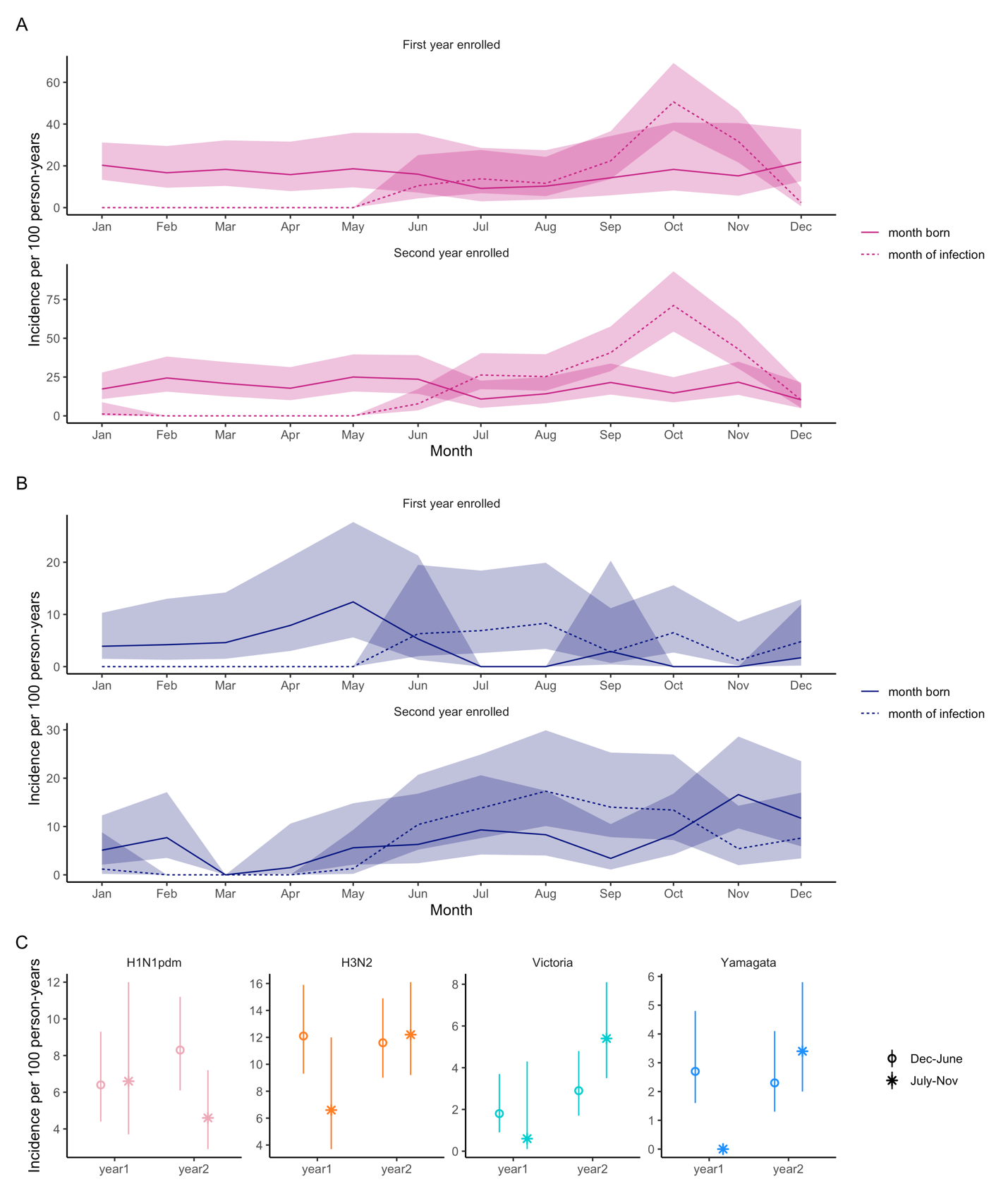

Figure S4. Symptomatic influenza incidence in the first and second years enrolled in infants enrolled in their first 6 months of life, by (sub)type and lineage. Panels A) and B) show incidence by month born (solid line) and month of infection (dotted line); shaded regions represent confidence interval. Panel C) shows incidence by month born categorized by months of primary influenza circulation, July-November compared to December-June, by subtype and lineage.

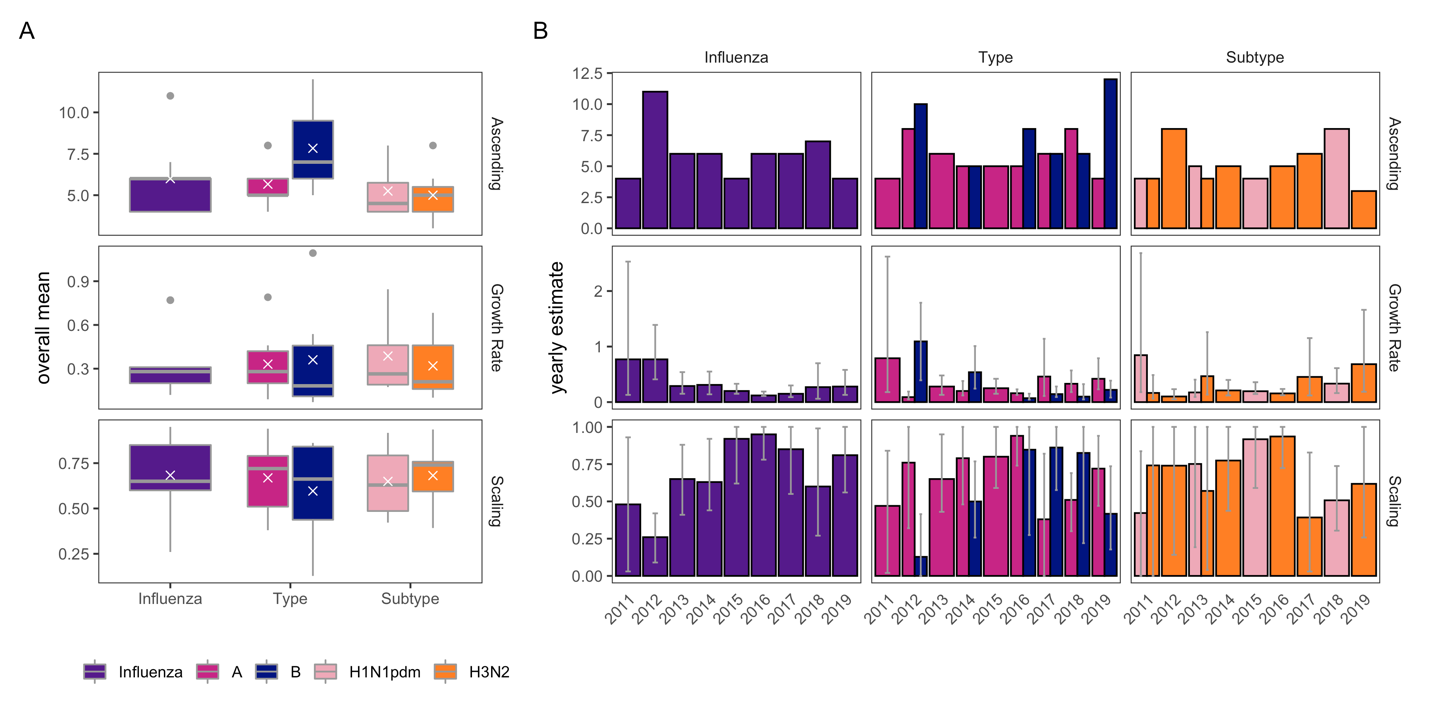

Figure S5. Other estimated transmission parameters: ascending phase, growth rate, and growth scaling factor. A) Overall transmission parameter means. B) Transmission parameter estimates by year. Confidence intervals are shown for growth rates and scaling factors; ascending period is the time from epidemic start to peak.

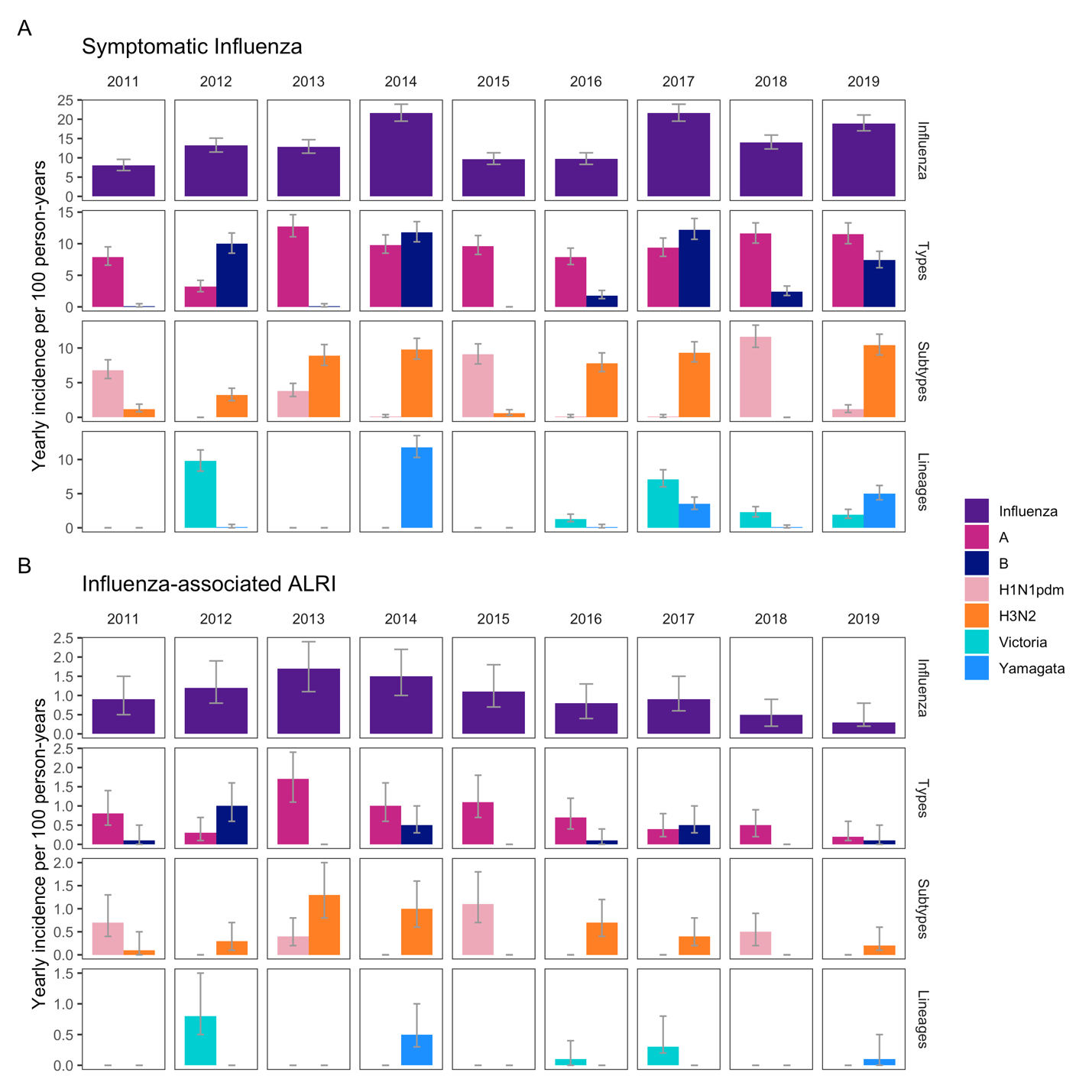

Figure S6. Incidence of symptomatic influenza and influenza-associated ALRI in the Nicaraguan Pediatric Influenza Cohort Study (NPICS), 2011-2019, by year.

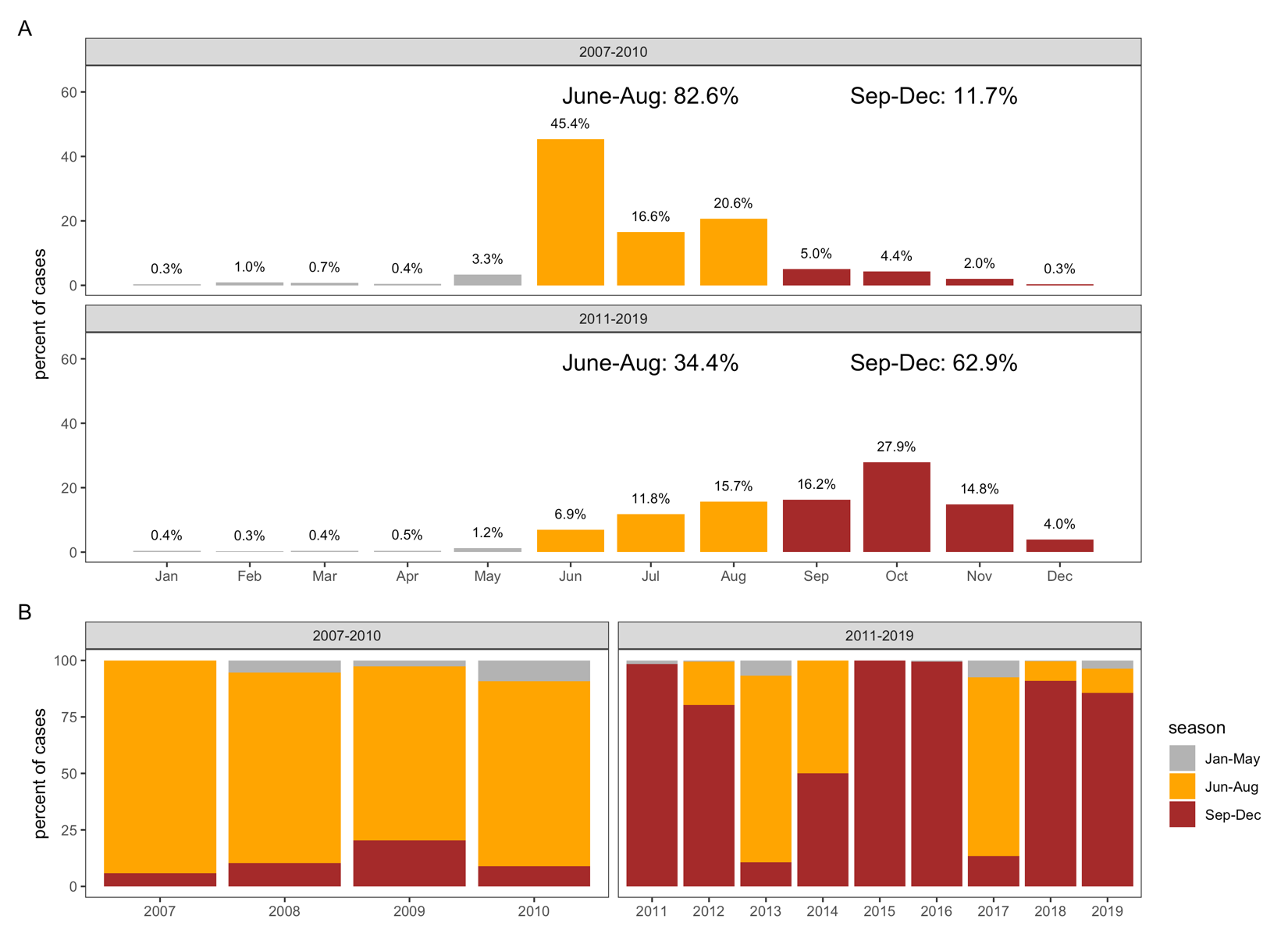

Figure S7. Influenza epidemic timing

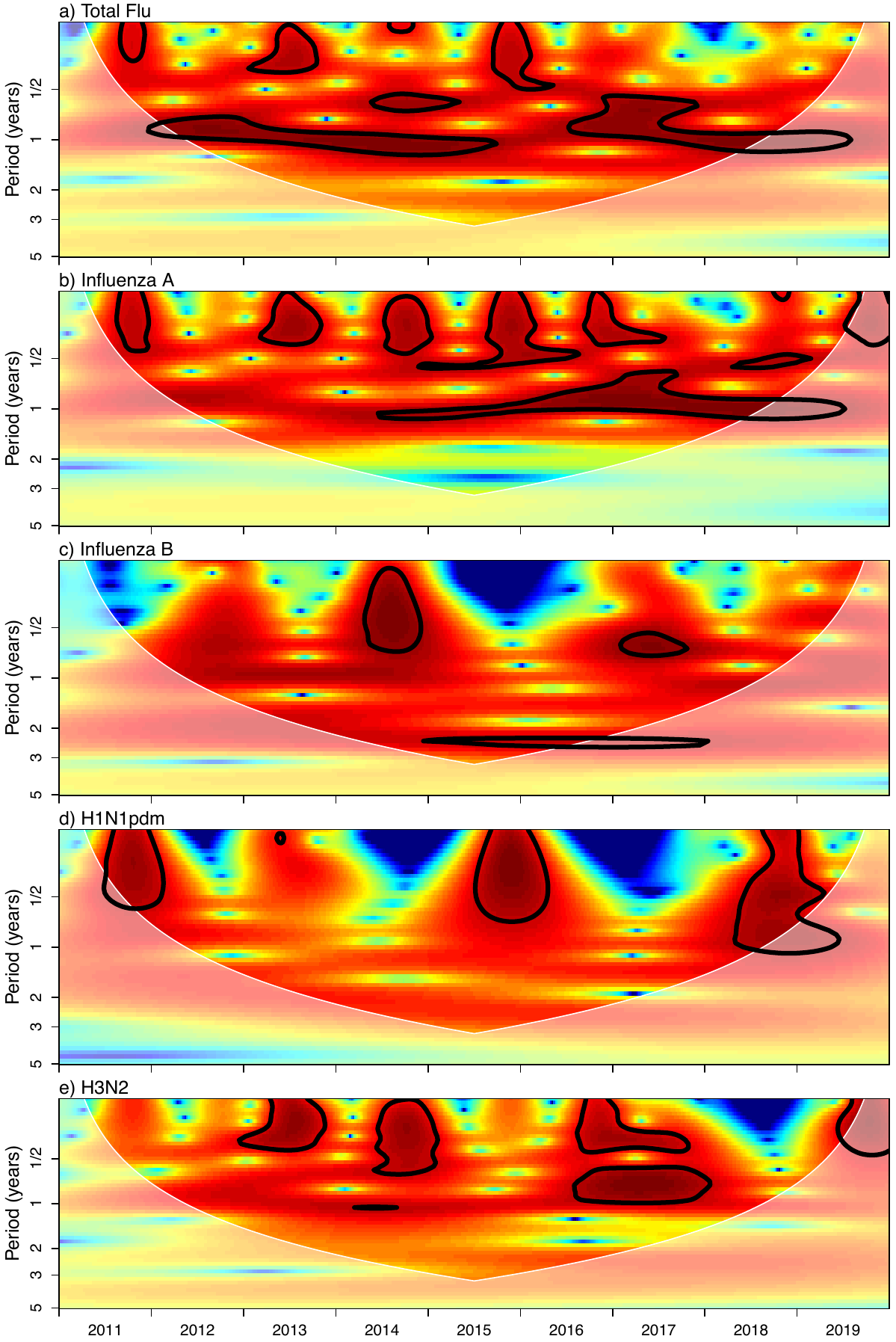

Figure S8. Influenza seasonality by Wavelet analysis. Wavelet analyses of log transformed + 1 data for: a) all symptomatic influenza, where an annual dominant period with significance can be seen within the cone of influence, b) H1N1, where dominant periods exist at the annual and biannual periods without significance, c) H3N2, where an annual dominant period can be seen, though significance is only present for one year, d) Influenza B, which reveals a 2.5 year dominant period with significance and e) Influenza A, where an annual dominant period with significance can be observed.

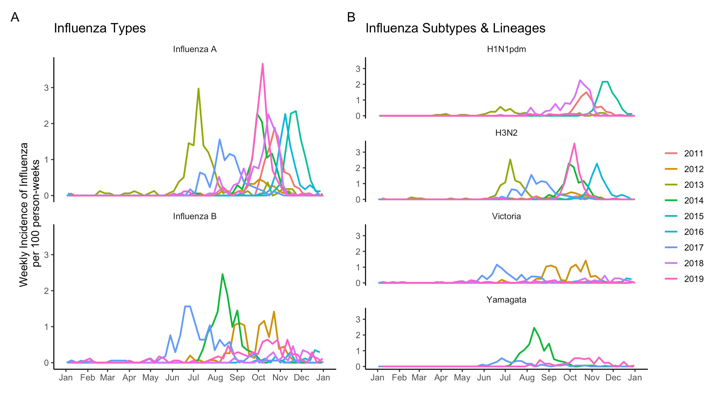

Figure S9. Yearly influenza epidemics, by month. A) types, and B) subtypes and lineages.
